# Performance evaluation of the LumiraDx SARS-CoV-2 & Flu A/B Test in diagnosing COVID-19 and influenza in patients with respiratory symptoms

**DOI:** 10.1101/2022.07.20.22277845

**Authors:** Jayne Ellis, Poppy Guest, Vicki Lawson, Julia Loecherbach, Nigel Lindner, Andrew McCulloch

## Abstract

**Introduction:** Coronavirus disease 2019 (COVID-19) and influenza share similar symptoms, which hampers diagnosis. Given that they require different containment and treatment strategies, fast and accurate distinction between the two infections is needed. This study evaluates the sensitivity and specificity of the LumiraDx SARS-CoV-2 & Flu A/B Test for simultaneous detection of severe acute respiratory syndrome coronavirus 2 (SARS-CoV-2) and influenza A/B from a single nasal swab.

**Methods:** Nasal samples were collected from patients as part of the ASPIRE (NCT04557046) and INSPIRE (NCT04288921) studies at point-of-care testing sites in the USA. ASPIRE study participants were included after developing COVID-19 symptoms in the last 14 days or following a positive SARS-CoV-2 test in the last 48 hours. INSPIRE study participants were included after developing influenza symptoms in the last 4 days. Samples were extracted into proprietary buffer and analysed using the LumiraDx SARS-CoV-2 & Flu A/B Test. A reference sample was taken from each subject, placed into universal transport medium and tested using reference SARS-CoV-2 and influenza reverse transcription polymerase chain reaction (RT-PCR) tests. The test and reference samples were compared using the positive percent agreement (PPA) and negative percent agreement (NPA), together with their 95% confidence intervals (CI).

**Results:** Analysis of the data from the ASPIRE (N=124) and INSPIRE (N=159) studies revealed high levels of agreement between the LumiraDx SARS-CoV-2 & Flu A/B Test and the reference tests in detecting SARS-CoV-2 (PPA=95.5% [95% CI: 84.9%, 98.7%]; NPA=96.0% [95% CI: 90.9%, 98.3%]), influenza A (PPA=83.3% [95% CI: 66.4%, 92.7%]; NPA=97.7% [95% CI: 93.4%, 99.2%]) and influenza B (PPA=80.0% [95% CI: 62.7%, 90.5%]; NPA=95.3% [95% CI: 90.2%, 97.9%]).

**Conclusions:** The LumiraDx SARS-CoV-2 & Flu A/B Test shows a high agreement with the reference RT-PCR tests while simultaneously detecting and differentiating between SARS-CoV-2 and influenza A/B.

Trial registration, ClinicalTrials.gov identifier: NCT04557046 and NCT04288921

## Introduction

The virus that causes coronavirus disease 2019 (COVID-19) in humans, known as the severe acute respiratory syndrome coronavirus 2 (SARS-CoV-2), emerged towards the end of 2019.[1] In most people who are infected with SARS-CoV-2, symptoms develop within 4–14 days of incubation and vary in severity. The most common symptoms include fever, dry cough and fatigue. The most severe symptoms are pneumonia and low blood oxygen levels, which can be fatal.[2, 3] According to the World Health Organization (WHO), as of 31^st^ January 2022, over 5.5 million global deaths have been attributed to COVID-19.[4] A timely and accurate diagnosis of COVID-19 is essential to help reduce SARS-CoV-2 transmission, and to protect individuals at risk of hospitalisation and fatality. Currently, reverse transcription polymerase chain reaction (RT-PCR) tests are the most accurate method of testing, but they are time- and resource-intensive.[5, 6]

Influenza viruses are also known to cause infectious respiratory diseases in humans and these can be mistaken for COVID-19 owing to the similarity in symptoms: cough, fever, fatigue and muscle pain.[7, 8] However, unlike COVID-19, the symptoms of influenza usually arise within 1–4 days following an infection.[9] While influenza is considered to be less threatening to the public than COVID-19, with 0.29–0.65 million annual deaths globally according to the WHO,[10] it can be severe and life-threatening in children, the elderly and people suffering with other conditions.[11] Influenza A is the most common type of influenza virus; it mutates quickly and is responsible for the majority of influenza outbreaks.[8] The second most common type of influenza virus is influenza B. Although influenza B is less infectious, its severity and mortality rate are comparable to those of type A.[12]

As the symptoms of COVID-19 and influenza are similar,[7] there is a need for a single-swab SARS-CoV-2 and influenza A/B combination test that can distinguish between these infections. This could facilitate appropriate clinical responses from healthcare professionals to target treatment and containment effectively.[13] While global influenza levels have been low since the COVID-19 pandemic started, infections are likely to rise as governments remove their COVID-19 related restrictions, allowing infectious respiratory diseases to spread more easily within the population. A rapid diagnostic combination test could facilitate fast differentiation between SARS-CoV-2 and influenza infections.[14, 15] The Academy of Medical Sciences report, *COVID-19: Preparing for the Future*, recommends integrating combined multiplex testing, to distinguish between SARS-CoV-2 and influenza, into primary and community care settings to reduce the transmission of both viruses.[16]

The highly sensitive SARS-CoV-2 and influenza A/B RT-PCR tests currently available on the market, including combined multiplex RT-PCR assays, have relatively long turnaround times, are expensive and require laboratory resources.[17-21] The LumiraDx SARS-CoV-2 & Flu A/B Test has been developed to rapidly, accurately and simultaneously test for SARS-CoV-2 and influenza A/B infections from a single nasal swab sample at the point of care.[22] This study evaluates the clinical performance of the rapid microfluidic immunofluorescence LumiraDx SARS-CoV-2 & Flu A/B Test.

## Methods

### Study design

Owing to the lack of circulating influenza since the start of the COVID-19 pandemic, the performance of the LumiraDx SARS-CoV-2 & Flu A/B Test was evaluated using frozen samples, previously collected via the prospective ASPIRE (NCT04557046) and INSPIRE (NCT04288921) clinical studies. Paired anterior nasal swabs for investigative and reference tests were collected from participants of the ASPIRE and INSPIRE studies using Nasal FLOQSwabs^®^-502CS01 (Copan Diagnostics Inc., Murrieta, CA, USA). Frozen samples collected under the ASPIRE and INSPIRE protocols were transported to LumiraDx (Stirling, UK). At the LumiraDx site, the test samples were thawed for a minimum of 30 minutes at room temperature, stirred and tested within 2 hours on the LumiraDx SARS-CoV-2 & Flu A/B Test in line with the manufacturer’s instructions for use.[23] Samples were kept cold using ice during testing. The reference and test samples were blinded from the operators to avoid any bias.

The ASPIRE study was conducted across six point-of-care testing sites in the USA. Participants of the ASPIRE study were of any age and sex, and were included following the onset of COVID-19 symptoms in the last 14 days or following a positive SARS-CoV-2 test within the past 48 hours. COVID-19 symptoms were defined as one or more of the following: fever, cough, shortness of breath, difficulty breathing, muscle pain, headache, sore throat, chills, repeated shaking with chills, new loss of taste and/or smell, congestion, runny nose, diarrhoea, nausea and vomiting. Samples were consecutively collected from participants between 26^th^ June 2020 and 24^th^ September 2020. One test and one reference anterior nasal swab were collected at the same time from each participant using the dual nares sampling method. The test samples were placed in the LumiraDx Extraction Buffer, frozen within 1 hour and stored at −20°C, before being transported at ≤ −70°C to LumiraDx, where they were processed in line with the manufacturer’s instructions for use. The reference samples were placed in 3 mL BD Universal Viral Transport Medium (Franklin Lakes, NJ, USA). The reference samples were transferred under refrigerated conditions for testing at the TriCore Reference Laboratories (Albuquerque, NM, USA), where they were processed and tested on the Roche cobas^®^ 6800 SARS-CoV-2 systems (Basel, Switzerland). Reference RT-PCR results were considered positive when the cycle threshold (Ct) was <33.

The INSPIRE study was carried out during the 2019–2020 influenza / respiratory syncytial virus (RSV) season in the USA across 13 point-of-care testing sites. Participants of the INSPIRE study were of any age and sex, and were included following the development of influenza symptoms within the last 4 days. Influenza symptoms were defined as fever and at least one of the following: stuffy or runny nose, sneezing, cough, sore throat, dyspnoea, wheezing, fatigue, weakness and/or malaise, arthralgia, myalgia, anorexia, vomiting, diarrhoea or headache. Samples were consecutively collected between 6^th^ January 2020 and 2^nd^ March 2020. One test and one reference anterior nasal swab were taken from both nostrils of each participant. The order in which samples were collected was randomised for the reference and test swabs to avoid bias in sampling. The test samples were frozen within 1 hour in the LumiraDx Extraction Buffer and stored at −20°C before being transported at ≤ −70°C to LumiraDx. The reference samples were placed into 3 mL BD Universal Viral Transport Medium and transported at 4°C to the TriCore Reference Laboratories, where they were tested on the Cepheid Xpert^®^ Xpress Flu/RSV assay (Sunnyvale, CA, USA).

### The investigational LumiraDx SARS-CoV-2 & Flu A/B Test

The LumiraDx SARS-CoV-2 & Flu A/B Test is a rapid microfluidic immunofluorescence assay that uses separate channels for SARS-CoV-2-, influenza A- and influenza B-specific assays. It is designed to detect SARS-CoV-2, influenza A and/or influenza B nucleocapsid proteins (NPs) from a single nasal swab sample. The antibodies on the test strip form particle–particle complexes with the nasal sample NPs. Upon binding, fluorescence is emitted in a concentration-dependent manner and detected by the testing device; the presence of SARS-CoV-2 and/or influenza A/B is reported on the instrument touch screen as ‘positive’ or ‘negative’ for each analyte. The test provides simultaneous and specific detection of SARS-CoV-2 and influenza A/B with a 12-minute turnaround time. Moreover, it contains built-in quality controls, which include an automated test strip expiration date verification and sample volume checks prior to the running of the test. The testing device can monitor many elements: electrical component operation; heater operation; battery charge state; mechanical actuators and sensors; test strip performance; controls; and optical system performance.

### Statistical analysis

These studies were set up in compliance with the US Food and Drug Administration (FDA) Antigen Template for Test Developers.[24] Statistical analyses were conducted using Microsoft Excel (Redmond, WA, USA) to determine positive percent agreement (PPA), negative percent agreement (NPA), positive predictive value (PPV), negative predictive value (NPV) and overall percent agreement, together with their associated two-sided Wilson score 95% confidence intervals (CIs) for the LumiraDx SARS-CoV-2 & Flu A/B Test in comparison to reference RT-PCR test results. Only test samples with available reference samples were analysed. The sensitivity of the LumiraDx SARS-CoV-2 & Flu A/B Test in symptomatic participants was assessed against an acceptance criteria of ≥80% agreement with the reference RT-PCR, based on the FDA Emergency Use Authorization (EUA) Antigen Template for Test Developers.[24]

### Ethical approval

Ethical approval was received from the WIRB-Copernicus Group (WCG) Institutional Review Board (IRB) for the ASPIRE study under protocol number CS-1211-01 (WCG IRB 20201775). Ethical approval was received from the WCG IRB for the INSPIRE study under protocol numbers CS-LUMFLURSV19-01 (CS-1176-01; WCG IRB 20193352) and CS-LUMFLURSV19-01A (CS-1176-01A; WCG IRB 20193211). Written consent from adult participants and legal parents/guardians of minor participants was obtained for study participation and publication.

## Results

### Study population

A total of 288 patients were recruited for the ASPIRE and INSPIRE studies. Of these, 283 subject samples were included in the final data analysis: 124 were from ASPIRE (SARS-CoV-2-positive and -negative cases) and 159 were from INSPIRE (Influenza A and B-positive and -negative cases) (**Figure 1**). Five subjects were excluded from these studies: two from the ASPIRE study (days since symptom onset >12 days or unknown) and three from the INSPIRE study (instrument malfunction). Using reference RT-PCR test results, the percentage prevalence values of SARS-CoV-2, influenza A and influenza B in the overall study population were calculated as 26.2%, 18.9% and 18.9%, respectively. The prevalence of SARS-CoV-2, influenza A and influenza B across age groups, identified by the LumiraDx device in agreement with the reference RT-PCR test, showed that SARS-CoV-2 was most prevalent in the 22–59 and >60 (years) age groups, whereas influenza A and B was most prevalent in the ≤ 5 and 6–21 (years) age groups (**Supplementary table 1**).

**Figure 1:**
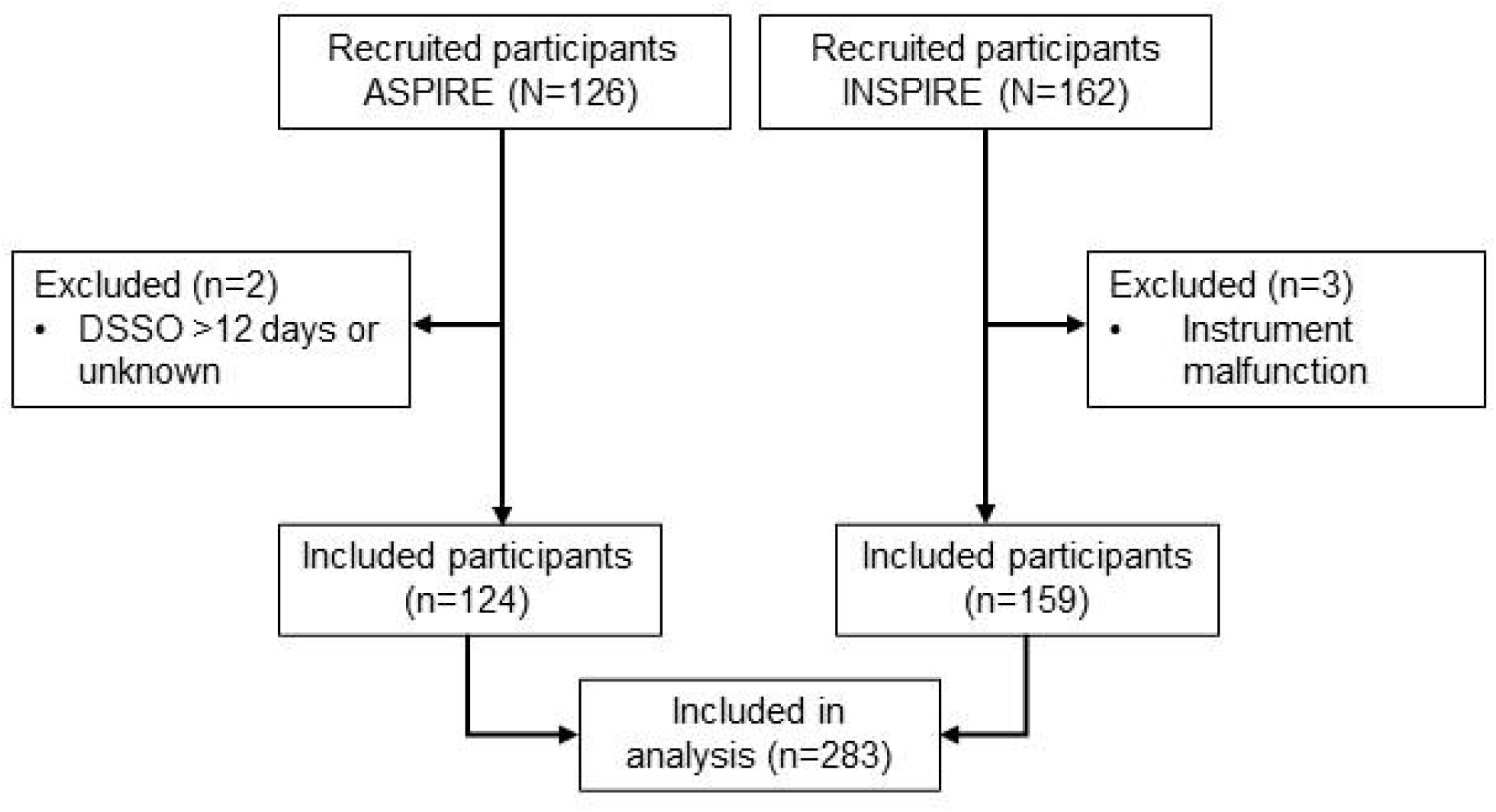
Flow of participants in ASPIRE and INSPIRE studies DSSO, days since symptom onset

### SARS-CoV-2 and influenza A/B antigen assay validation

Of the 168 samples in the SARS-CoV-2 analysis with confirmed RT-PCR results from both INSPIRE and ASPIRE studies, 44 tested positive by RT-PCR and 42 tested positive in agreement with the reference using the LumiraDx SARS-CoV-2 & Flu A/B Test, which resulted in a PPA of 95.5% (95% CI: 84.9%, 98.7%) and a PPV of 89.4% (95% CI: 77.4%, 95.4%) (**Table 1**).

**Table 1:**
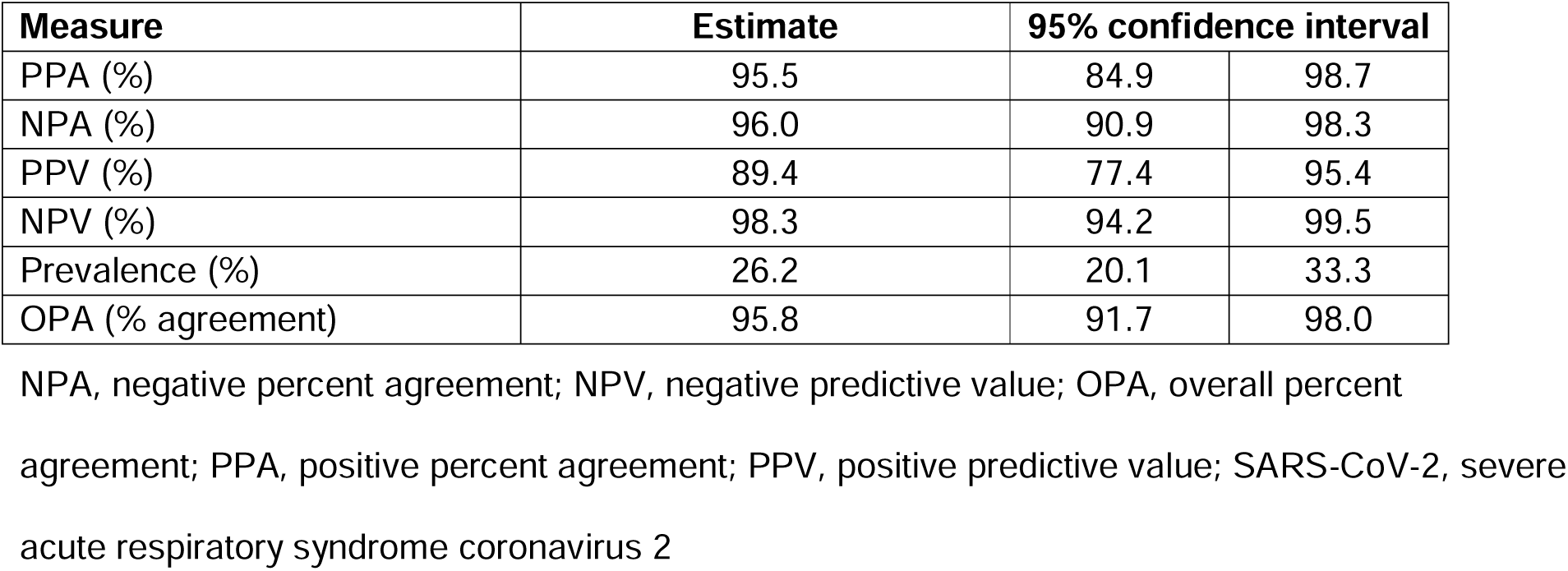
SARS-CoV-2 detection performance measures for the LumiraDx SARS-CoV-2 & Flu A/B Test

| Measure | Estimate | 95% confidence interval |  |
| --- | --- | --- | --- |
| PPA (%) | 95.5 | 84.9 | 98.7 |
| NPA (%) | 96.0 | 90.9 | 98.3 |
| PPV (%) | 89.4 | 77.4 | 95.4 |
| NPV (%) | 98.3 | 94.2 | 99.5 |
| Prevalence (%) | 26.2 | 20.1 | 33.3 |
| OPA (% agreement) | 95.8 | 91.7 | 98.0 |

NPA, negative percent agreement; NPV, negative predictive value; OPA, overall percent agreement; PPA, positive percent agreement; PPV, positive predictive value; SARS-CoV-2, severe acute respiratory syndrome coronavirus 2

Furthermore, from 124 samples that tested negative by RT-PCR, 119 were confirmed by the LumiraDx SARS-CoV-2 & Flu A/B Test, resulting in an NPA of 96.0% (95% CI: 90.9%, 98.3%) and an NPV of 98.3% (95% CI: 94.2%, 99.5%) (**Table 1**). The highest agreement between the LumiraDx SARS-CoV-2 & Flu A/B Test and RT-PCR was found in samples with Ct <30 (**Table 2**). Further stratification based on days since symptom onset revealed that the highest agreement was found in samples collected within 3 days of symptom onset (**Figure 2**).

**Table 2:**
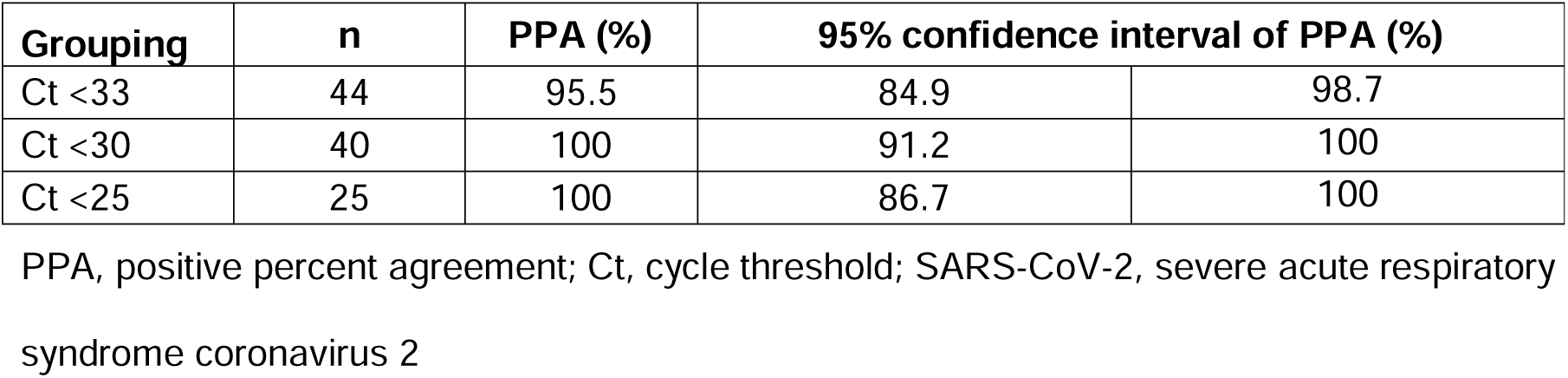
SARS-CoV-2 detection sensitivity of the LumiraDx SARS-CoV-2 & Flu A/B Test across cycle threshold groupings

| Grouping | n | PPA (%) | 95% confidence interval of PPA (%) |  |
| --- | --- | --- | --- | --- |
| Ct <33 | 44 | 95.5 | 84.9 | 98.7 |
| Ct <30 | 40 | 100 | 91.2 | 100 |
| Ct <25 | 25 | 100 | 86.7 | 100 |

PPA, positive percent agreement; Ct, cycle threshold; SARS-CoV-2, severe acute respiratory syndrome coronavirus 2

**Figure 2:**
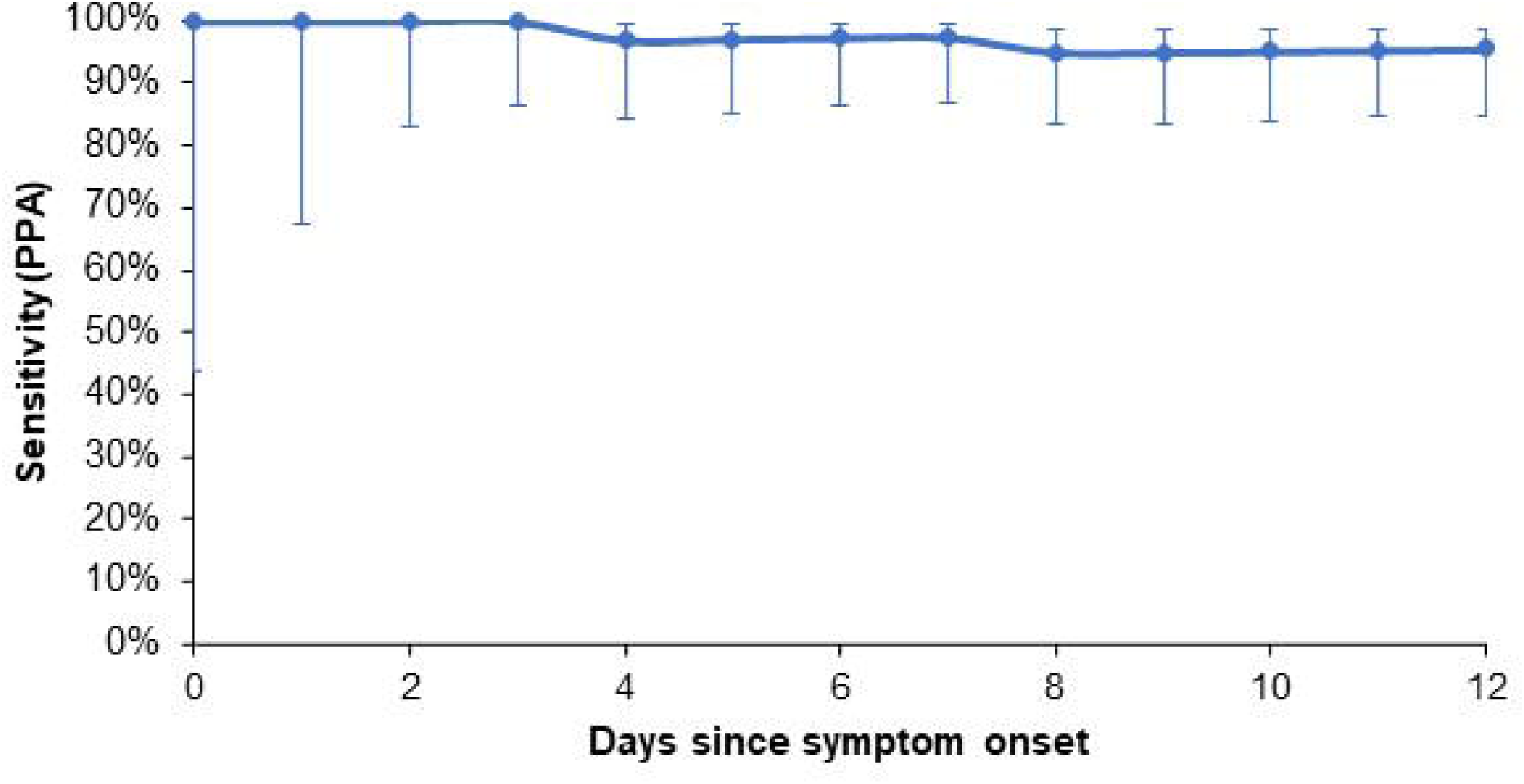
SARS-CoV-2 detection sensitivity of the LumiraDx SARS-CoV-2 & Flu A/B Test across days since symptom onset, with cycle threshold <33 PPA, positive percent agreement

Overall, 30 samples tested positive and 129 samples tested negative using RT-PCR for influenza A during the INSPIRE study. Of these samples, 25 tested positive and 126 tested negative, respectively, using the LumiraDx SARS-CoV-2 & Flu A/B Test. This resulted in a PPA and PPV of 83.3% (95% CI: 66.4%, 92.7%) and 89.3% (95% CI: 72.8%, 96.3%), respectively, and an NPA and NPV of 97.7% (95% CI: 93.4%, 99.2%) and 96.2% (95% CI: 91.4%, 98.4%), respectively (**Table 3**).

**Table 3:** Influenza A detection performance measures for the LumiraDx SARS-CoV-2 & Flu A/B Test

| Measure | Estimate | 95% confidence interval |  |
| --- | --- | --- | --- |
| PPA (%) | 83.3 | 66.4 | 92.7 |
| NPA (%) | 97.7 | 93.4 | 99.2 |
| PPV (%) | 89.3 | 72.8 | 96.3 |
| NPV (%) | 96.2 | 91.4 | 98.4 |
| Prevalence (%) | 18.9 | 13.5 | 25.7 |
| OPA (% agreement) | 95.0 | 90.4 | 97.4 |

NPA, negative percent agreement; NPV, negative predictive value; OPA, overall percent agreement; PPA, positive percent agreement; PPV, positive predictive value; SARS-CoV-2, severe acute respiratory syndrome coronavirus 2

In addition, 30 samples tested positive and 129 samples tested negative using RT-PCR for influenza B during the INSPIRE study. Of these, 24 were confirmed positive and 123 were confirmed negative, respectively, using the LumiraDx SARS-CoV-2 & Flu A/B Test. Analysis revealed a PPA of 80.0% (95% CI: 62.7%, 90.5%) and an NPA of 95.3% (95% CI: 90.2%, 97.9%).Furthermore, a PPV of 80.0% (95% CI: 62.7%, 90.5%) and an NPV of 95.3% (95% CI: 90.2%, 97.9%) were obtained (**Table 4**).

**Table 4:** Influenza B detection performance measures for the LumiraDx SARS-CoV-2 & Flu A/B Test

| Measure | Estimate | 95% confidence interval |  |
| --- | --- | --- | --- |
| PPA (%) | 80.0 | 62.7 | 90.5 |
| NPA (%) | 95.3 | 90.2 | 97.9 |
| PPV (%) | 80.0 | 62.7 | 90.5 |
| NPV (%) | 95.3 | 90.2 | 97.9 |
| Prevalence (%) | 18.9 | 13.5 | 25.7 |
| OPA (% agreement) | 92.5 | 87.3 | 95.6 |

NPA, negative percent agreement; NPV, negative predictive value; OPA, overall percent agreement; PPA, positive percent agreement; PPV, positive predictive value; SARS-CoV-2, severe acute respiratory syndrome coronavirus 2

## Discussion

The COVID-19 pandemic continues to evolve globally, and while vaccination and testing strategies have been widely implemented to deal with the emergency, the focus has now shifted to learning to live with the virus. This is challenging because respiratory infections such as COVID-19 and influenza have overlapping symptoms.[16] Therefore, there is a need for point-of-care multiplex testing for COVID-19 and influenza, using simple nasal swab specimens, would facilitate timely diagnosis, treatment and isolation decisions. Such testing could be deployed in hospitals, primary care, care homes and community pharmacies.[16]

This study evaluated the clinical performance of the LumiraDx SARS-CoV-2 & Flu A/B Test using retrospective samples. The results demonstrated a high degree of positive and negative agreement between the LumiraDx SARS-CoV-2 & Flu A/B Test and RT-PCR. The sensitivity of the test remained high for SARS-CoV-2 up to 12 days since symptom onset. Moreover, a similar sensitivity to the reference SARS-CoV-2 EUA RT-PCR test was detected in sample groups with Ct <30, which suggests the test can detect relatively low viral loads.

In comparison to the currently available multiplex SARS-CoV-2 and Flu A/B PCR tests, the LumiraDx SARS-CoV-2 & Flu A/B Test offers some advantages.[17-21] For example, the average turnaround time is reduced from a few hours to 12 minutes.[21] Additionally, the test is portable and can be used at the point of care such as community test centres. In reference to the currently marketed SARS-CoV-2 antigen tests with an average sensitivity of 71.2%, the LumiraDx SARS-CoV-2 & Flu A/B Test was notably superior with a PPA of 95.5%.[25] The sensitivity of the LumiraDx SARS-CoV-2 & Flu A/B Test in this study was similar to those reported for the LumiraDx SARS-CoV-2 Antigen Test in previous studies in symptomatic (97.5%) and asymptomatic (82.1%) study populations.[26, 27] The LumiraDx SARS-CoV-2 Antigen Test uses the same antibody reagents as the LumiraDx SARS-CoV-2 & Flu A/B Test. Currently available multiplex SARS-CoV-2 & Flu A/B antigen point-of-care tests have SARS-CoV-2 sensitivities ranging from 86.7%–95.2%.[28]

The sensitivity of the LumiraDx SARS-CoV-2 & Flu A/B Test compared with the RT PCR test was 83.3% for influenza A and 80% for influenza B; these values are higher than the average of 69% reported for currently available influenza A and B antigen tests.[29] These improved sensitivities, when compared with other influenza antigen tests utilising conventional lateral flow technology, may be due to the microfluidic immunofluorescence technology used in this test system. This will be further evaluated in a prospective study during the next seasonal influenza outbreak.

The main study limitation was the use of retrospective remnant samples, owing to a lack of circulating influenza when the study was conducted. The lack of circulating influenza is thought to be due to COVID-19 pandemic preventive measures, such as mask-wearing and physical distancing. Sensitivity of influenza antigen assays has been reported to be unaffected by freezing and thawing of samples, which indicates that the use of frozen samples in our study did not influence results.[30] This will need to be demonstrated in a prospective study during the next influenza season. Another limitation in this study was the size of the cohort; however, the small study size was aligned with the FDA EUA antigen template for new diagnostic tests.[31] The template allows smaller study samples owing to the COVID-19 pandemic emergency. Future prospective studies will need to examine larger patient cohorts. Another potential setback of the study is that SARS-CoV-2 samples collected are likely to be of the wild-type form as they were obtained between June and September 2020.[32] However, the LumiraDx SARS-CoV-2 Antigen Test shows excellent performance against all reported variants, including Omicron.[33] Hence, SARS-CoV-2 variants are not expected to affect test performance.

## Conclusion

The LumiraDx SARS-CoV-2 & Flu A/B Test offers a highly sensitive single nasal swab test for the detection and differentiation between SARS-CoV-2 and influenza A/B viruses at the point of care. The test provides rapid results and removes the need for lengthy additional tests to differentiate between the infections that present with similar symptoms but require different containment and treatment strategies. The LumiraDx SARS-CoV-2 & Flu A/B Test can potentially allow healthcare professionals to quickly and specifically identify the infection and decide on appropriate containment and treatment strategies. Multiplex testing for respiratory infections in point-of-care settings is essential as seasonal influenza and COVID-19 outbreaks may become the new normal.[34]

## Supporting information

STARD checklist

## Data Availability

All data produced in the present work are contained in the manuscript

## Acknowledgements

The authors acknowledge Norbert Chmurzynski from imc (integrated medhealth communication) for medical writing support, funded by LumiraDx, and Dr Stephen Young from the TriCore Reference Laboratories for his contributions to data collection. The authors would like to thank participating patients for their involvement.

## Authorship confirmation

All authors met all four requirements for authorship as outlined by International Committee of Medical Journal Editors (ICMJE). All authors read and approved the final version of the manuscript.

## Data availability

All data generated or analysed during this study are included in this published article as supplementary information files.

## Tables

**Supplementary table 1:** Positive subjects identified by the LumiraDx SARS-CoV-2 & Flu A/B Test in agreement with the reference reverse transcription polymerase chain reaction tests across age groups

| **Assay** | **Age (years)** | **n** | **Positive** | **Prevalence (%)** |
| --- | --- | --- | --- | --- |
| SARS-CoV-2 | ≤5 | 49 | 0 | 0.0 |
|  | 6–21 | 114 | 5 | 4.4 |
|  | 22–59 | 157 | 34 | 21.7 |
|  | ≥60 | 27 | 3 | 11.1 |
| Influenza A | ≤5 | 49 | 5 | 10.2 |
|  | 6–21 | 114 | 13 | 11.4 |
|  | 22–59 | 157 | 7 | 4.5 |
|  | ≥60 | 27 | 0 | 0.0 |
| Influenza B | ≤5 | 49 | 5 | 10.2 |
|  | 6–21 | 114 | 17 | 14.9 |
|  | 22–59 | 157 | 2 | 1.3 |
|  | ≥60 | 27 | 0 | 0.0 |

SARS-CoV-2, severe acute respiratory syndrome coronavirus 2

