## Supplementary material for "Performance evaluation of the LumiraDx SARS-CoV-2 & Flu A/B Test in diagnosing COVID-19 and influenza in patients with respiratory symptoms": STARD checklist

|  | **Section & Topic** | **No** | **Item** | **Reported on page #** |
| --- | --- | --- | --- | --- |
|  | **TITLE OR ABSTRACT** |  |  | 1 |
|  |  | **1** | Identification as a study of diagnostic accuracy using at least one measure of accuracy  (such as sensitivity, specificity, predictive values, or AUC) | 2 |
|  | **ABSTRACT** |  |  | 2-3 |
|  |  | **2** | Structured summary of study design, methods, results, and conclusions  (for specific guidance, see STARD for Abstracts) | 2-3 |
|  | **INTRODUCTION** |  |  | 4-5 |
|  |  | **3** | Scientific and clinical background, including the intended use and clinical role of the index test | 4-5 |
|  |  | **4** | Study objectives and hypotheses | 5 |
|  | **METHODS** |  |  | 5-8 |
|  | *Study design* | **5** | Whether data collection was planned before the index test and reference standard  were performed (prospective study) or after (retrospective study) | 5 |
|  | *Participants* | **6** | Eligibility criteria | 6 |
|  |  | **7** | On what basis potentially eligible participants were identified  (such as symptoms, results from previous tests, inclusion in registry) | 6 |
|  |  | **8** | Where and when potentially eligible participants were identified (setting, location and dates) | 6 |
|  |  | **9** | Whether participants formed a consecutive, random or convenience series | 6 |
|  | *Test methods* | **10a** | Index test, in sufficient detail to allow replication | 7 |
|  |  | **10b** | Reference standard, in sufficient detail to allow replication | 5-7 |
|  |  | **11** | Rationale for choosing the reference standard (if alternatives exist) | 4(introduction) |
|  |  | **12a** | Definition of and rationale for test positivity cut-offs or result categories  of the index test, distinguishing pre-specified from exploratory | 6 |
|  |  | **12b** | Definition of and rationale for test positivity cut-offs or result categories  of the reference standard, distinguishing pre-specified from exploratory | 6 |
|  |  | **13a** | Whether clinical information and reference standard results were available  to the performers/readers of the index test | 6 |
|  |  | **13b** | Whether clinical information and index test results were available  to the assessors of the reference standard | 6-7 |
|  | *Analysis* | **14** | Methods for estimating or comparing measures of diagnostic accuracy | 7-8 |
|  |  | **15** | How indeterminate index test or reference standard results were handled | 7-8 |
|  |  | **16** | How missing data on the index test and reference standard were handled | 7-8 |
|  |  | **17** | Any analyses of variability in diagnostic accuracy, distinguishing pre-specified from exploratory | 7-8 |
|  |  | **18** | Intended sample size and how it was determined | 8 |
|  | **RESULTS** |  |  | 9-10 |
|  | *Participants* | **19** | Flow of participants, using a diagram | Figure 1 |
|  |  | **20** | Baseline demographic and clinical characteristics of participants | Not available |
|  |  | **21a** | Distribution of severity of disease in those with the target condition | 8 and Supplementary table 1 and |
|  |  | **21b** | Distribution of alternative diagnoses in those without the target condition | Not available |
|  |  | **22** | Time interval and any clinical interventions between index test and reference standard | 5,6 |
|  | *Test results* | **23** | Cross tabulation of the index test results (or their distribution)  by the results of the reference standard | Tables 1-4 |
|  |  | **24** | Estimates of diagnostic accuracy and their precision (such as 95% confidence intervals) | Tables 1-4 |
|  |  | **25** | Any adverse events from performing the index test or the reference standard | Not applicable |
|  | **DISCUSSION** |  |  | 10-12 |
|  |  | **26** | Study limitations, including sources of potential bias, statistical uncertainty, and generalisability | 11-12 |
|  |  | **27** | Implications for practice, including the intended use and clinical role of the index test | 10,12 |
|  | **OTHER INFORMATION** |  |  |  |
|  |  | **28** | Registration number and name of registry | 2 |
|  |  | **29** | Where the full study protocol can be accessed | Not available |
|  |  | **30** | Sources of funding and other support; role of funders | 11 |

STARD 2015

### AIM

STARD stands for “Standards for Reporting Diagnostic accuracy studies”. This list of items was developed to contribute to the completeness and transparency of reporting of diagnostic accuracy studies. Authors can use the list to write informative study reports. Editors and peer-reviewers can use it to evaluate whether the information has been included in manuscripts submitted for publication.

### Explanation

A **diagnostic accuracy study** evaluates the ability of one or more medical tests to correctly classify study participants as having a **target condition.** This can be a disease, a disease stage, response or benefit from therapy, or an event or condition in the future. A medical test can be an imaging procedure, a laboratory test, elements from history and physical examination, a combination of these, or any other method for collecting information about the current health status of a patient.

The test whose accuracy is evaluated is called **index test.** A study can evaluate the accuracy of one or more index tests. Evaluating the ability of a medical test to correctly classify patients is typically done by comparing the distribution of the index test results with those of the **reference standard**. The reference standard is the best available method for establishing the presence or absence of the target condition. An accuracy study can rely on one or more reference standards.

If test results are categorized as either positive or negative, the cross tabulation of the index test results against those of the reference standard can be used to estimate the **sensitivity** of the index test (the proportion of participants *with* the target condition who have a positive index test), and its **specificity** (the proportion *without* the target condition who have a negative index test). From this cross tabulation (sometimes referred to as the contingency or “2x2” table), several other accuracy statistics can be estimated, such as the positive and negative **predictive values** of the test. Confidence intervals around estimates of accuracy can then be calculated to quantify the statistical **precision** of the measurements.

If the index test results can take more than two values, categorization of test results as positive or negative requires a **test positivity cut-off**. When multiple such cut-offs can be defined, authors can report a receiver operating characteristic (ROC) curve which graphically represents the combination of sensitivity and specificity for each possible test positivity cut-off. The **area under the ROC curve** informs in a single numerical value about the overall diagnostic accuracy of the index test.

The **intended use** of a medical test can be diagnosis, screening, staging, monitoring, surveillance, prediction or prognosis. The **clinical role** of a test explains its position relative to existing tests in the clinical pathway. A replacement test, for example, replaces an existing test. A triage test is used before an existing test; an add-on test is used after an existing test.

Besides diagnostic accuracy, several other outcomes and statistics may be relevant in the evaluation of medical tests. Medical tests can also be used to classify patients for purposes other than diagnosis, such as staging or prognosis. The STARD list was not explicitly developed for these other outcomes, statistics, and study types, although most STARD items would still apply.

### DEVELOPMENT

This STARD list was released in 2015. The 30 items were identified by an international expert group of methodologists, researchers, and editors. The guiding principle in the development of STARD was to select items that, when reported, would help readers to judge the potential for bias in the study, to appraise the applicability of the study findings and the validity of conclusions and recommendations. The list represents an update of the first version, which was published in 2003.

More information can be found on [http://www.equator-network.org/reporting-guidelines/stard](http://www.equator-network.org/reporting-guidelines/stard/).
